# Macrophage expression and prognostic significance of the long pentraxin PTX3 in COVID-19

**DOI:** 10.1101/2020.06.26.20139923

**Authors:** Enrico Brunetta, Marco Folci, Barbara Bottazzi, Maria De Santis, Alessandro Protti, Sarah Mapelli, Roberto Leone, Ilaria My, Monica Bacci, Veronica Zanon, Gianmarco Spata, Andrea Gianatti, Marina Sironi, Claudio Angelini, Cecilia Garlanda, Michele Ciccarelli, Maurizio Cecconi, Alberto Mantovani

## Abstract

PTX3 is an essential component of humoral innate immunity, involved in resistance to selected pathogens and in the regulation of inflammation. PTX3 plasma levels are associated with poor outcome in systemic inflammatory conditions and vascular pathology. The present study was designed to assess expression and significance of PTX3 in COVID-19. By bioinformatics analysis of public databases PTX3 expression was detected in lung respiratory cell lines exposed to SARS-CoV-2. By analysis at single cell level of COVID-19 circulating mononuclear cells, we found that PTX3 was selectively expressed by monocytes among circulating leukocytes. Moreover, in lung bronchoalveolar lavage fluid, single cell analysis revealed selective expression of PTX3 in neutrophils and macrophages, which play a major role in the pathogenesis of the disease. By immunohistochemistry, PTX3 was expressed by lung myelomocytic cells, type 2 pneumocytes and vascular endothelial cells. PTX3 plasma levels were determined by ELISA in 96 consecutive patients with a laboratory-confirmed diagnosis of COVID-19. Higher PTX3 plasma levels were observed in 52 (54.2%) patients admitted in ICU (median 21.0ng/mL, IQT 15.5-46.3 vs 12.4ng/mL IQT 6.1-20.2 in ward patients; p=0.0017) and in 22 (23%) patients died by 28 days (39.8ng/mL, IQT 20.2-75.7 vs 15.7ng/mL, IQT 8.2-21.6 in survivors; p=0.0001). After determining an optimal PTX3 cut-off for the primary outcome, the Kaplan-Meier curve showed an increased mortality in patients with PTX3>22.25ng/mL (Log-rank tests p<0.0001). In Cox regression model, PTX3>22.25ng/mL showed an adjusted Hazard Ratio (aHR) of 7.6 (95%CI2.45-23.76) in predicting mortality. Performing a multivariate logistic regression including all inflammatory markers (PTX3, ferritin, D-Dimer, IL-6, and CRP), PTX3 was the only marker significantly associated with death (aHR 1.13;95%CI1.02-1.24; p=0.021). The results reported here suggest that circulating and lung myelomonocytic cells are a major source of PTX3 and that PTX3 plasma levels can serve as a strong prognostic indicator of short-term mortality in COVID-19.

## INTRODUCTION

Highly pathogenic betacoronaviruses, causing Severe Acute Respiratory Syndrome (SARS), Middle East Respiratory Syndrome (MERS), and the currently pandemic COVID-19, affect the lower respiratory tract leading to critical acute respiratory distress syndrome (ARDS) and fatality in a high percentage of cases ^1-4^. SARS-CoV-2 infection is characterized by variable clinical forms with symptoms including fever, cough, and general malaise in mild and moderate cases ^5^, which progress to severe pneumonia or ARDS, shock and/or multiple organ failure, requiring hospitalization in Intensive Care Units (ICU), in more severe cases. The high morbidity and mortality observed in COVID-19 pandemics are caused by alveolar damage and pneumonia ^6,7^, cardiovascular complications, and multiorgan failure ^8^.

SARS-CoV-2 interacts with ACE2 expressed by pneumocytes in the alveolar lining, leading to lung injury. ACE2 is also widely expressed on endothelial cells, thus possibly explaining the evidence of direct viral infection of the endothelium, diffuse endothelial inflammation, and widespread microvascular dysfunction, leading to organ ischaemia, inflammation, edema, and a procoagulant state ^4,9,10^. In addition, uncontrolled activation of innate and adaptive immunity in response to the infection results in hyperinflammatory responses, as demonstrated by the cytokine storm and activation of macrophages and neutrophils in COVID-19 patients, which, by affecting lung tissue and blood vessels, contribute to ARDS pathogenesis, shock and multiorgan failure ^4,7,11^.

PTX3 is a key component of humoral innate immunity, belonging to the family of pentraxins ^12^. In contrast with its relative, the short pentraxin C reactive protein (CRP), essentially produced by the liver in response to IL-6 during the acute phase response ^13^, PTX3 is rapidly produced by several cell types, including myeloid cells, endothelial cells, and respiratory epithelial cells, in particular in response to IL-1, TNFα, microbial molecules, and tissue damage ^12^. PTX3 is an essential component of humoral innate immunity, involved in resistance to selected pathogens and in the regulation of inflammation.

The similarity with CRP prompted investigations as to the usefulness of PTX3 as a marker in diverse human conditions of infective or inflammatory origin. The local production by different cell types at inflammatory sites and the release of the preformed protein by neutrophils in response to primary proinflammatory cytokines or microbial recognition accounts for the rapidity of PTX3 increase in these conditions. Increased PTX3 plasma concentrations were described in infections of fungal, bacterial and viral origin ^14-17^, severe inflammatory response syndrome (SIRS), sepsis ^18-21^, and cardiovascular diseases ^22-24^. In different pathological conditions high PTX3 plasma levels were associated with disease severity and mortality (e.g. ^18-20^). Moreover, PTX3 has been shown to serve as a biomarker of disease activity in inflammatory conditions involving the vascular bed, ranging from atherosclerosis to vasculitis (e.g.^22,25-30^).

Previous findings on the prognostic significance of PTX3 in systemic inflammatory conditions as well as in vascular pathology ^18,29^ prompted the present investigation which was designed to assess PTX3 expression and plasma levels in COVID-19. Here we report that, based on a bioinformatic analysis on public databases, PTX3 was strongly induced by SARS-COV-2 in respiratory tract epithelial cells. At single cell level, COVID-19 monocytes and lung macrophages expressed PTX3. Finally, PTX3 plasma levels were a strong predictor of 28-day mortality in hospitalized COVID-19 patients.

## Methods

### Bioinformatic analysis

Data relative to the transcriptional response to SARS-CoV-2 infection were derived from datasets deposited within the Gene Expression Omnibus (GEO). Data relative to bulk transcription in human normal bronchial (NHBE) and malignant cell lines (Calu-3 and A549) upon SARS-CoV-2 infection, were derived from the experiments within the series GSE147507 ^31^. Raw bulk RNA-Seq reads were quality inspected with the software “FastQC” (https://www.bioinformatics.babraham.ac.uk/projects/fastqc/) and aligned with STAR (version 2.6.1) ^32^ on the GRCh38 genome guided by GENCODE annotation (version 33).

Gene summarized counts were processed in R, genes whose expression was major than 2 reads were filtered and vst normalized with the R package DESeq2 ^33^. Significantly changing genes upon SARS-CoV-2 infection were obtained with DESeq2 by comparing each infected cell line with the respective mock-treated counterpart. Gene identifiers conversions were performed with the “org.Hs.eg.db” library (https://www.bioconductor.org/packages//2.10/data/annotation/html/org.Hs.eg.db.html). Plots were rendered with the R library “ggplot2” (https://ggplot2.tidyverse.org).

Available single cell RNA-Seq experiments, related to BALF of SARS-CoV2 individuals, were obtained from the public repositories Gene Expression Omnibus (GEO) and FigShare platform under the identifiers GSE145926 ^34,35^ and the FigShare platform (https://figshare.com/articles/COVID-19_severity_correlates_with_airway_epithelium-immune_cell_interactions_identified_by_single-cell_analysis/12436517). scRNA-Seq datasets of SARS-CoV-2 infected PBMC (deposited in GEO under the series GSE150728) ^36^ were explored with the portal cellxgene (https://chanzuckerberg.github.io/cellxgene/) and obtained from “The COVID19 Cell Atlas portal” (https://www.covid19cellatlas.org/#wilk20). Sparse count matrices or Seurat objects were obtained as released and processed with the R package “Seurat” ^37^ and confirmed with the published pipelines shared by respective authors.

Classification of clusters was performed according to the authors’ parameters and annotations were performed with the Blueprint-Encode signatures. The distribution of PTX3 expression was obtained after imputation with the “Rmagic” package ^38^.

### Immunohistochemistry

Immunohistochemical staining was performed as follows: 3-µm-thick sections were prepared from formalin-fixed paraffin-embedded autoptic tissue lung blocks from a COVID-19 patient and were dried in a 60°C for 20’. The sections were placed in a BOND-III Automated Immunohistochemistry Vision Biosystem (Leica Microsystems GmbH, Wetzlar, Germany) according to the following protocol. First, tissues were deparaffinized and pre-treated with the Epitope Retrieval Solution 1 (Citrate-buffer pH 5.9-6.1) at 100°C for 10 min. After washing steps, peroxidase blocking was carried out for 5 min using the Bond Polymer Refine Detection Kit DC9800 (Leica Microsystems GmbH). Tissues were again washed and then incubated with the primary antibody (affinity purified rabbit IgG anti-human PTX3) ^39^ for 15 min. Subsequently, tissues were incubated with polymer for 8 min and developed with DAB-Chromogen for 10 min.

### Study design and participants

This cohort study analyzed a cohort of 96 patients. We included all males and non-pregnant females, 18 years of age or older, admitted to Humanitas Clinical and Research Center (Rozzano, Milan, Italy) between March 4th and May 16rd, 2020 (data cutoff on May 13rd) with a laboratory-confirmed diagnosis of COVID-19. Hospital admission criteria were based on a positive assay for SARS-CoV-2 associated with respiratory failure requiring oxygen therapy, or radiological evidence of significant pulmonary infiltrates on chest computed tomography (CT) scan, or reduction in respiratory/cardiopulmonary reserve as assessed by 6 minutes walking test, or due to frailty related with patient comorbidity. We assessed an outcome of death. 52 patients of 96 (54%) were transferred to ICU because requiring invasive ventilation or non-invasive mechanical ventilation with oxygen fraction over 60%. Patients with continuous positive airway pressure therapy (CPAP) were followed up by ICU outreach team and ward physicians in COVID-19 wards. Acute respiratory syndrome (ARDS) was defined according to the Berlin definition ^40^.

### Laboratory test, demographic, and medical history

Laboratory testing at hospital admission included: complete blood count, renal and liver function (transaminase, total/direct/indirect bilirubin, gamma-glutamyl transferase, alkaline phosphatase), creatinine kinase, lactate dehydrogenase, myocardial enzymes, electrolytes and triglycerides. A panel of acute phase reactants including interleukin-6 (IL-6), serum ferritin, D-dimer, C-reactive protein (CRP), fibrinogen, and procalcitonin (PCT) was performed. Body temperature, blood pressure, heart rate, peripheral saturation, and respiratory rate were measured in all patients. Chest CT scan and arterial blood gas analysis were performed in the emergency department. In all patients PTX3 was measured within the first few days after the admission date (mean 2.1+1.6 days). Pneumococcal and Legionella urinary antigen tests were routinely performed. Nasopharyngeal swab for influenza A, B and H1N1 was also routinely performed to exclude co-infections. Additional microbiological tests were performed to exclude other pathogens as possible etiological agents when suggested by clinical conditions (bacterial cultures of sputum, blood and urine). We obtained a comprehensive present and past medical history from patients. Positivity was assessed on the basis of reverse-transcriptase– polymerase-chain-reaction (RT-PCR) assay for SARS-CoV-2 on a respiratory tract sample tested by our laboratory, in accordance with the protocol established by the WHO (https://www.who.int/emergencies/diseases/novel-coronavirus-2019/technical-guidance/laboratory-guidance). Due to the high false negative rate of RT-PCR from pharyngeal swab, two different swabs were performed in every patient to increase the detection rate ^41^. In cases of negative assay in throat-swab specimens, but with suggestive clinical manifestations, presence of contact history or suggestive radiological evidence for COVID-19, the detection was performed on bronchoalveolar lavage fluid (BAL) or endotracheal aspirate, which has higher diagnostic accuracy. All demographics, medical history and laboratory tests were extracted from electronic medical records and were checked by a team of three expert physicians. The study was approved by the local Ethical Committee (authorization 233/20), and the requirement for informed consent was waived.

### Sample collection and PTX3 measurement

Venous blood samples were collected during the first 5 days after hospital admission (mean 2.1± standard deviation 1.6 days), centrifuged, and EDTA plasma was stored at -80°C until use. PTX3 plasma levels were measured, as previously described ^20^, by a sandwich ELISA (detection limit 0.1 ng/mL, inter-assay variability from 8 to 10%) developed in-house, by personnel blind to patients’ characteristics. In each analytical session a sample obtained from a pool of plasma from healthy donors was used as internal control. The mean PTX3 concentration measured in this sample was 1.88±0.6 ng/mL.

### Statistical methods

Demographic, clinical, laboratory, and outcome data were obtained from electronic medical records and patient chart notes using a standardized data collection form. Descriptive statistics included means with standard deviations (SD) and medians with interquartile ranges (IQR) for continuous variables, and frequency analyses (percentages) for categorical variables. Wilcoxon rank-sum tests were applied to continuous variables and two-tailed Fisher’s exact tests were used for categorical variables. Linearity of continuous variables was checked by comparing models with the linear term to the model with restricted cubic splines. The optimal cut-off levels of PTX3 for predicting 28-day outcome of death have been investigated using receiver operating characteristic (ROC) curves. The correlation between variables was evaluated by Spearman’s rank correlation coefficient (rho). To identify the association between PTX3 levels and the outcome in hospitalized COVID-19 patients, we used time-to-event (survival) methods for censored observations. The composite study endpoint was “death” within 28 days from hospital admission. Time to event was defined as the time from hospital admission until the date of event or censoring. Patients discharged early and alive from the hospital were considered event-free through day 28 ^42^. May 13rd 2020 was considered as data cutoff. Kaplan–Meier estimates were used to draw the cumulative incidence curves, compared by log-rank tests. Furthermore, multivariable Cox proportional hazards (PH) models of prognostic factors were used. The analyses were based on non-missing data (missing data not imputed). Confounders were selected according to a review of the literature, statistical relevance, and consensus opinion by an expert group of physicians and methodologists. After fitting the model, the PH assumption was examined on the basis of Schoenfeld residuals. The hazard ratios (HR) were presented with their 95% confidence intervals (CI) and the respective p-values. A ratio higher than 1.0 implies a higher probability of event compared to the reference group.

## Results

### PTX3 expression is induced in COVID-19

We conducted an *in silico* bioinformatic analysis of the expression of PTX3 using public databases. As shown in Figure 1A SARS-CoV-2 strongly induced or amplified PTX3 transcript expression in three lines representative of respiratory tract epithelial cells (Calu-3; A549; human normal bronchial cells, NHBE) (dataset GSE147507) ^31^. Bulk RNA-seq of purified monocytes (not shown) and bioinformatic analysis at single cell level of peripheral blood monononuclear cells obtained from COVID-19 patients, revealed that PTX3 was selectively expressed by COVID-19 monocytes (dataset GSE150728) ^36^ (Figure 1B). Interestingly, at single cell level CD16 monocytes were negative for PTX3 expression (not shown). Moreover, bioinformatic analysis at single cell level of COVID-19 bronchoalveolar lavage cells ^43^ revealed that PTX3 was strongly expressed in neutrophils and monocyte-macrophage populations, as identified by molecular signatures (Figure 1C, D). In the same dataset, epithelial cells in the fluid were negative.

**Figure 1.**
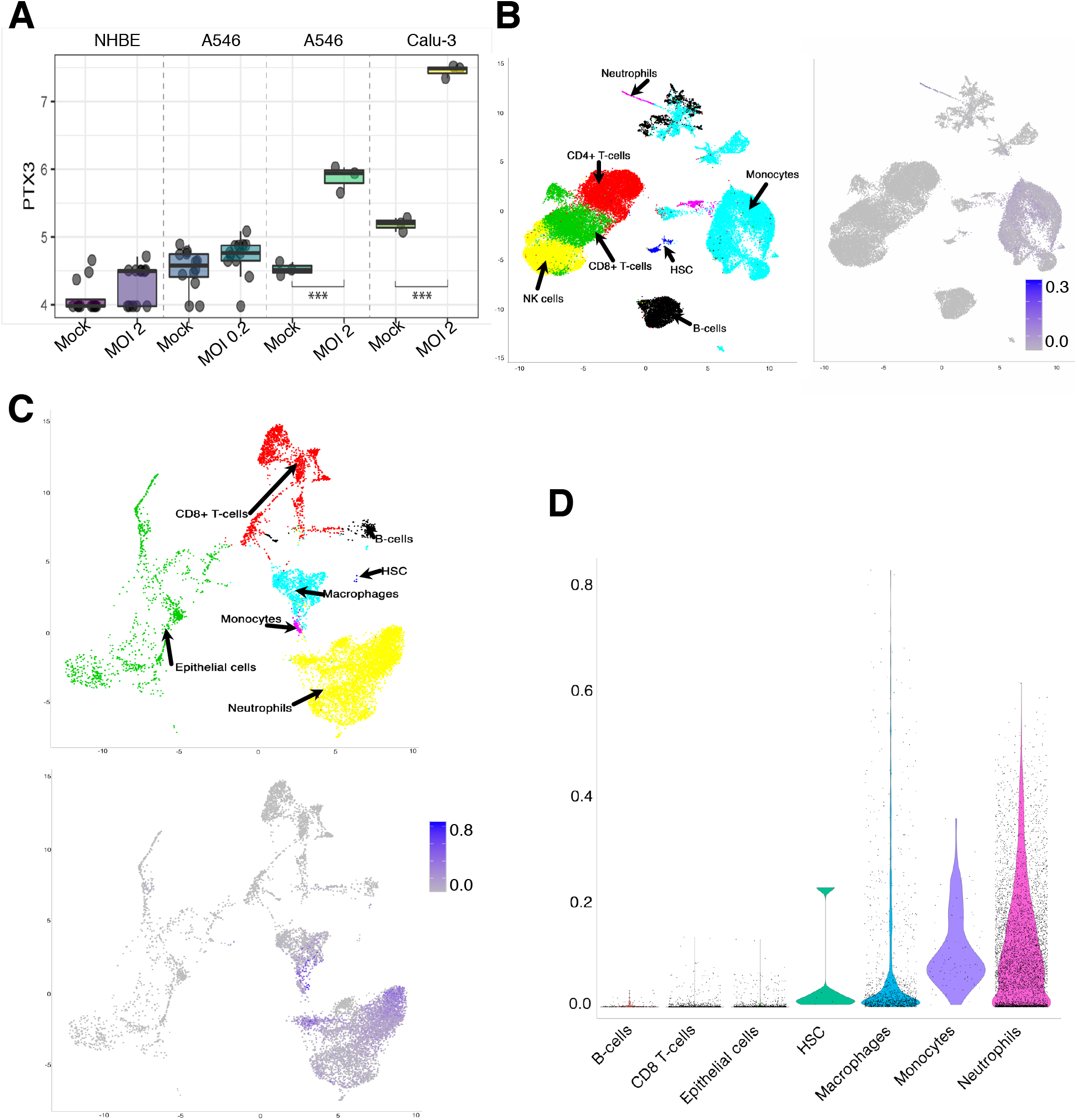
In silico analysis of PTX3 expression in SARS-CoV-2 infected cells and COVID-19 patients. A) PTX3 expression in *in vitro* SARS-CoV-2 infected respiratory epithelial cells: normal human bronchial epithelial cells -NHBC (at MOI 2) and human lung cancer cell lines A549 (at MOI 0.2 and 2) and Calu-3 (at MOI 2). B) Single cell RNA-seq of COVID-19 PBMC. Right panel, Uniform Manifold Approximation and Projection (UMAP) map, showing Seurat-guided clustering of PBMC populations. Each point represents a single-cell colored according to cluster designation. Left panel: Heatmap showing PTX3 expression. C) Single cell RNA-seq of COVID-19 BALF. Upper panel, UMAP showing Seurat-guided clustering of BALF populations. Lower panel: Heatmap showing PTX3 expression. D) Violin plots representing expression and distribution of PTX3 within predicted BALF populations. Expression values are relieved after Rmagic imputation.

Further confirmation of PTX3 expression in COVID-19 patients was obtained by immunohistochemistry on autopsy lung samples from subjects with a confirmed positivity to SARS-CoV-2. Figure 2A reports a typical staining on lung specimens showing PTX3 positivity in alveolar macrophages, as well as in the adventitia of small vessels and in type II pneumocytes. PTX3 immunostaining was also observed in multinucleated macrophages (Figure 2A). Interestingly, PTX3 staining was particularly intense in the adventitia and endothelium of small vessels occluded by a thrombus (Figure 2B).

**Figure 2.**
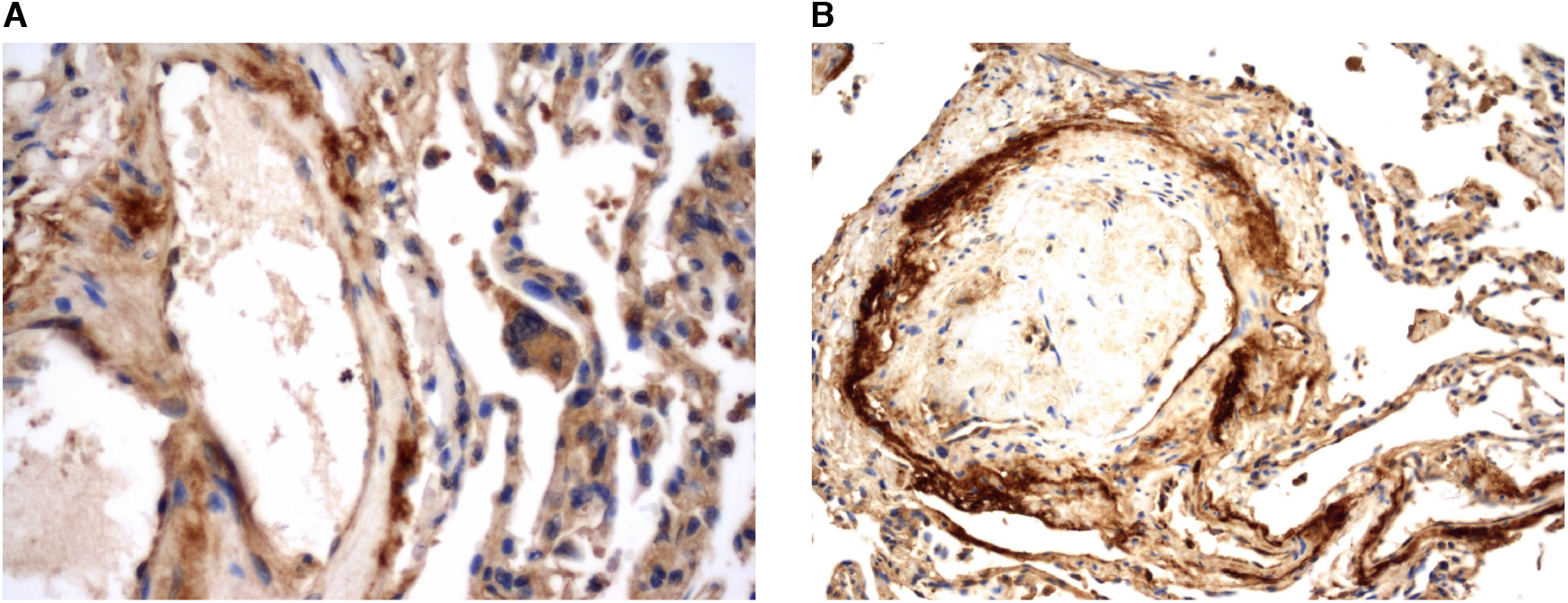
PTX3 immunolocalisation in COVID-19 lung specimens. A) PTX3 in the adventitia of small vessels, type II pneumocytes and alveolar macrophages. Note the granular cytoplasmic positivity in a multinuclear macrophage (immunoperoxidase stain, 40X magnification). B) PTX3 in the adventitia and endothelium of a small vessel occluded by a thrombus (immunoperoxidase stain, 20X magnification).

### Prognostic significance of PTX3 blood levels

Based on previous studies on the prognostic significance of PTX3 in inflammatory conditions and on transcript expression mainly in myelomonocytic cells, which play a key role in pathogenesis, we assessed PTX3 plasma levels in 96 consecutive COVID-19 patients admitted to the Humanitas Clinical and Research Center.

Demographic, clinical, and laboratory features of patients were shown in Table 1. All variables were also stratified by clinical outcome, dead versus alive (Table 1). In the overall population, patients had elevated PTX3 (mean 28.7± 29.8 ng/mL; median 17.3 ng/mL, IQT 10.0-39.8 ng/mL), CRP (mean 14.2±10.3 mg/dL; median 12.29 mg/dL, IQT 6.22-22.0 mg/dL), and IL-6 levels (median 48 pg/ml; IQT 21-115 pg/ml). Similarly to our previous study ^44^, we also observed elevated levels of ferritin (mean 634±608 µg/mL; median 536 µg/mL, IQT 151-822 µg/mL), D-dimer (mean 2404±7448 µg/mL; median 741 µg/mL, IQT 476-1456 µg/mL), and LDH (mean 371±168 U/L; median 345 U/L, IQT 268-424 U/L). A reduction in lymphocytes (mean 0.86±0.43 ×10^3^/mL; median 0.8 ×10^3^/mL, IQT 0.55-1.1 ×10^3^/mL) and eosinophils (mean 0.06±0.111 ×10^3^/mL; median 0 ×10^3^/mL, IQT 0-0.1 ×10^3^/mL) were also found, as previously reported ^44^.

**Table 1.**
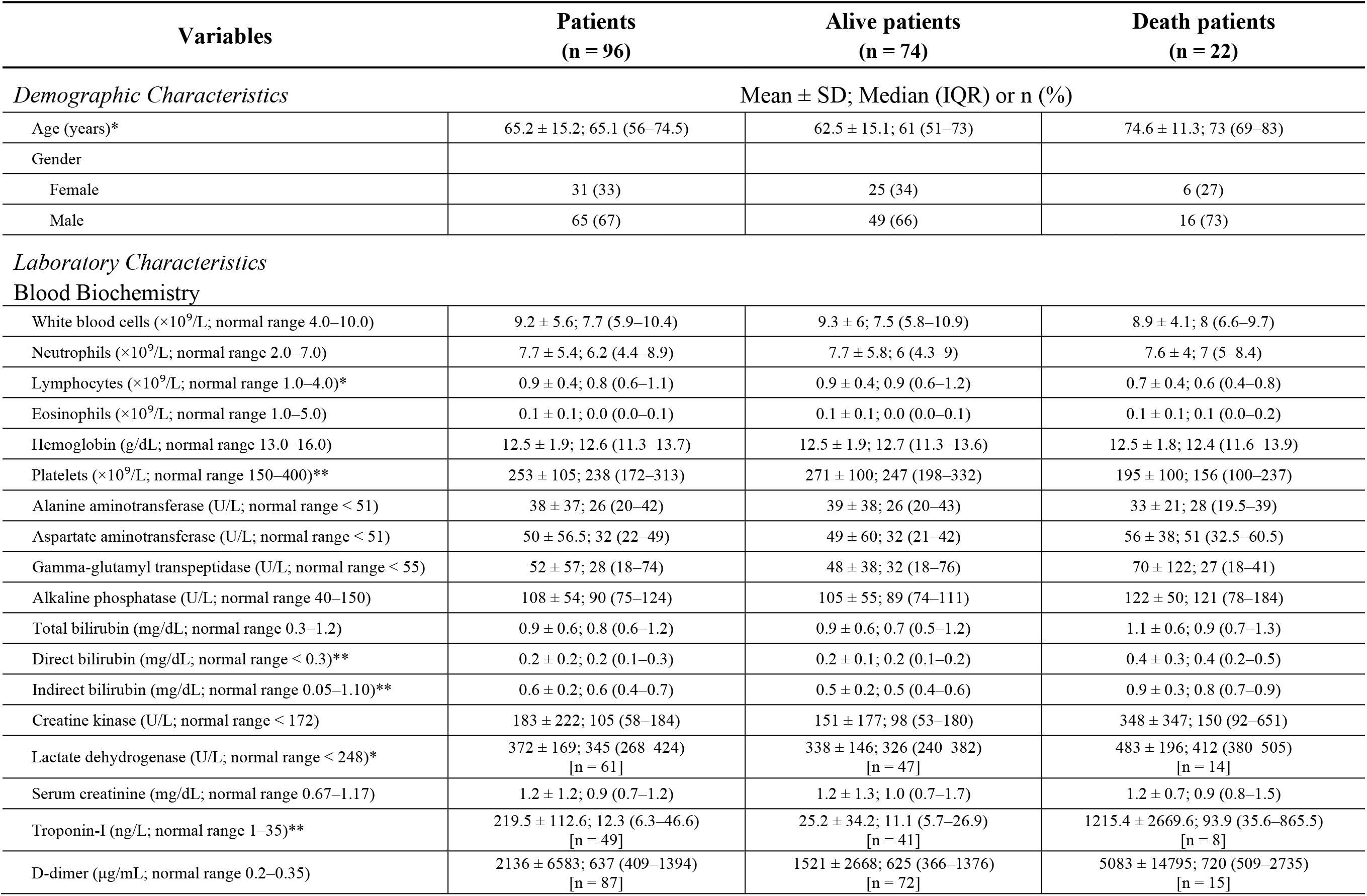
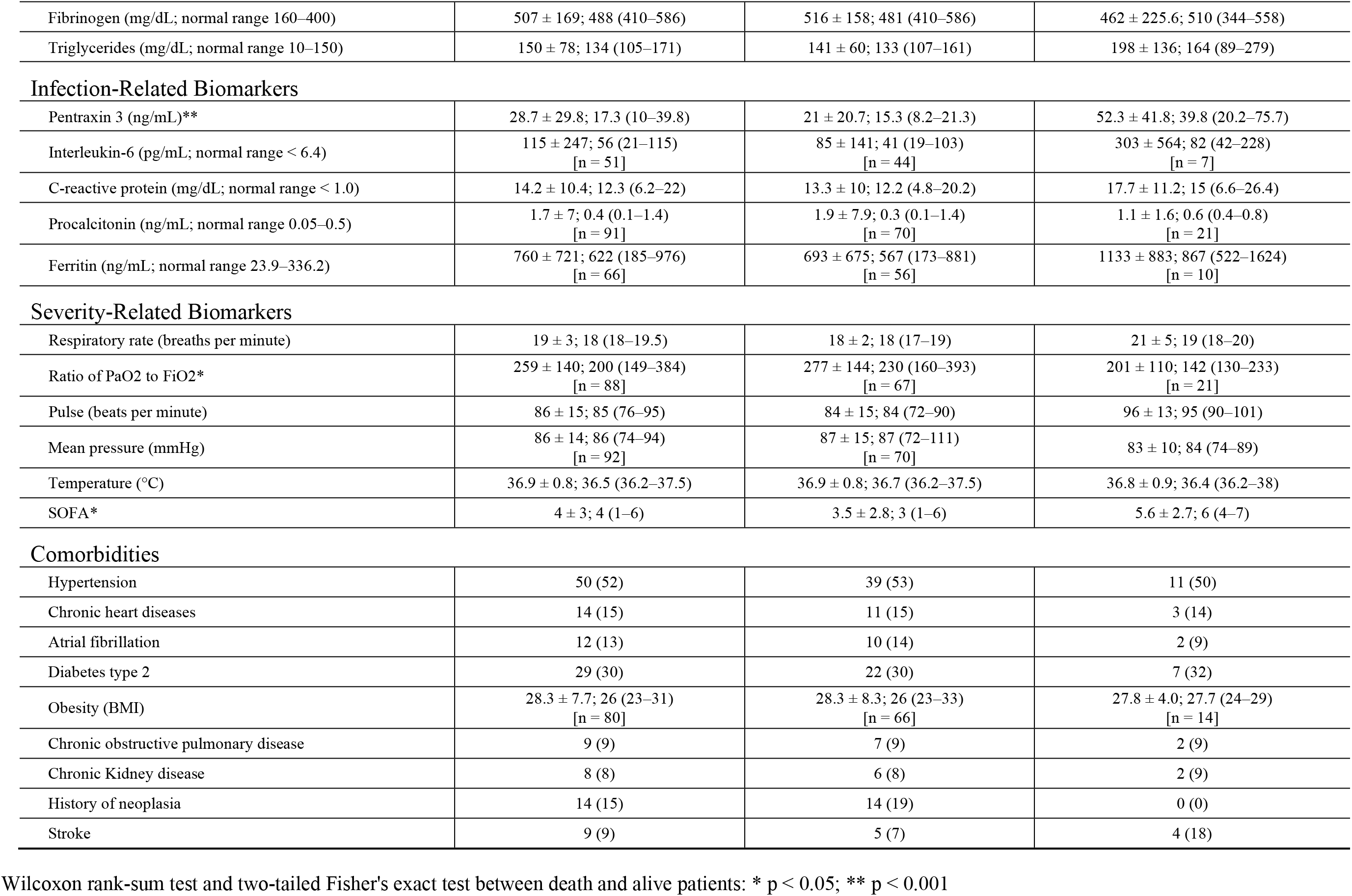
Demographics, laboratory and clinical characteristics of COVID-19 hospitalized patients grouped by different outcome.

| Variables | Patients<br>(n = 96) | Alive patients<br>(n = 74) | Death patients<br>(n = 22) |
| --- | --- | --- | --- |
| *Demographic Characteristics* |  |  |  |
| | Mean ± SD; Median (IQR) or n (%) | | |
| Age (years)* | 65.2 ± 15.2; 65.1 (56–74.5) | 62.5 ± 15.1; 61 (51–73) | 74.6 ± 11.3; 73 (69–83) |
| Gender |  |  |  |
| Female | 31 (33) | 25 (34) | 6 (27) |
| Male | 65 (67) | 49 (66) | 16 (73) |
| *Laboratory Characteristics* |  |  |  |
| **Blood Biochemistry** |  |  |  |
| White blood cells ($\times 10^9/L$; normal range 4.0–10.0) | 9.2 ± 5.6; 7.7 (5.9–10.4) | 9.3 ± 6; 7.5 (5.8–10.9) | 8.9 ± 4.1; 8 (6.6–9.7) |
| Neutrophils ($\times 10^9/L$; normal range 2.0–7.0) | 7.7 ± 5.4; 6.2 (4.4–8.9) | 7.7 ± 5.8; 6 (4.3–9) | 7.6 ± 4; 7 (5–8.4) |
| Lymphocytes ($\times 10^9/L$; normal range 1.0–4.0)* | 0.9 ± 0.4; 0.8 (0.6–1.1) | 0.9 ± 0.4; 0.9 (0.6–1.2) | 0.7 ± 0.4; 0.6 (0.4–0.8) |
| Eosinophils ($\times 10^9/L$; normal range 1.0–5.0) | 0.1 ± 0.1; 0.0 (0.0–0.1) | 0.1 ± 0.1; 0.0 (0.0–0.1) | 0.1 ± 0.1; 0.1 (0.0–0.2) |
| Hemoglobin (g/dL; normal range 13.0–16.0) | 12.5 ± 1.9; 12.6 (11.3–13.7) | 12.5 ± 1.9; 12.7 (11.3–13.6) | 12.5 ± 1.8; 12.4 (11.6–13.9) |
| Platelets ($\times 10^9/L$; normal range 150–400)** | 253 ± 105; 238 (172–313) | 271 ± 100; 247 (198–332) | 195 ± 100; 156 (100–237) |
| Alanine aminotransferase (U/L; normal range < 51) | 38 ± 37; 26 (20–42) | 39 ± 38; 26 (20–43) | 33 ± 21; 28 (19.5–39) |
| Aspartate aminotransferase (U/L; normal range < 51) | 50 ± 56.5; 32 (22–49) | 49 ± 60; 32 (21–42) | 56 ± 38; 51 (32.5–60.5) |
| Gamma-glutamyl transpeptidase (U/L; normal range < 55) | 52 ± 57; 28 (18–74) | 48 ± 38; 32 (18–76) | 70 ± 122; 27 (18–41) |
| Alkaline phosphatase (U/L; normal range 40–150) | 108 ± 54; 90 (75–124) | 105 ± 55; 89 (74–111) | 122 ± 50; 121 (78–184) |
| Total bilirubin (mg/dL; normal range 0.3–1.2) | 0.9 ± 0.6; 0.8 (0.6–1.2) | 0.9 ± 0.6; 0.7 (0.5–1.2) | 1.1 ± 0.6; 0.9 (0.7–1.3) |
| Direct bilirubin (mg/dL; normal range < 0.3)** | 0.2 ± 0.2; 0.2 (0.1–0.3) | 0.2 ± 0.1; 0.2 (0.1–0.2) | 0.4 ± 0.3; 0.4 (0.2–0.5) |
| Indirect bilirubin (mg/dL; normal range 0.05–1.10)** | 0.6 ± 0.2; 0.6 (0.4–0.7) | 0.5 ± 0.2; 0.5 (0.4–0.6) | 0.9 ± 0.3; 0.8 (0.7–0.9) |
| Creatine kinase (U/L; normal range < 172) | 183 ± 222; 105 (58–184) | 151 ± 177; 98 (53–180) | 348 ± 347; 150 (92–651) |
| Lactate dehydrogenase (U/L; normal range < 248)* | 372 ± 169; 345 (268–424)<br>[n = 61] | 338 ± 146; 326 (240–382)<br>[n = 47] | 483 ± 196; 412 (380–505)<br>[n = 14] |
| Serum creatinine (mg/dL; normal range 0.67–1.17) | 1.2 ± 1.2; 0.9 (0.7–1.2) | 1.2 ± 1.3; 1.0 (0.7–1.7) | 1.2 ± 0.7; 0.9 (0.8–1.5) |
| Troponin-I (ng/L; normal range 1–35)** | 219.5 ± 112.6; 12.3 (6.3–46.6)<br>[n = 49] | 25.2 ± 34.2; 11.1 (5.7–26.9)<br>[n = 41] | 1215.4 ± 2669.6; 93.9 (35.6–865.5)<br>[n = 8] |
| D-dimer ($\mu g/mL$; normal range 0.2–0.35) | 2136 ± 6583; 637 (409–1394)<br>[n = 87] | 1521 ± 2668; 625 (366–1376)<br>[n = 72] | 5083 ± 14795; 720 (509–2735)<br>[n = 15] |
| Fibrinogen (mg/dL; normal range 160–400) | 507 ± 169; 488 (410–586) | 516 ± 158; 481 (410–586) | 462 ± 225.6; 510 (344–558) |
| Triglycerides (mg/dL; normal range 10–150) | 150 ± 78; 134 (105–171) | 141 ± 60; 133 (107–161) | 198 ± 136; 164 (89–279) |
| Infection-Related Biomarkers |  |  |  |
| Pentraxin 3 (ng/mL)** | 28.7 ± 29.8; 17.3 (10–39.8) | 21 ± 20.7; 15.3 (8.2–21.3) | 52.3 ± 41.8; 39.8 (20.2–75.7) |
| Interleukin-6 (pg/mL; normal range < 6.4) | 115 ± 247; 56 (21–115)<br>[n = 51] | 85 ± 141; 41 (19–103)<br>[n = 44] | 303 ± 564; 82 (42–228)<br>[n = 7] |
| C-reactive protein (mg/dL; normal range < 1.0) | 14.2 ± 10.4; 12.3 (6.2–22) | 13.3 ± 10; 12.2 (4.8–20.2) | 17.7 ± 11.2; 15 (6.6–26.4) |
| Procalcitonin (ng/mL; normal range 0.05–0.5) | 1.7 ± 7; 0.4 (0.1–1.4)<br>[n = 91] | 1.9 ± 7.9; 0.3 (0.1–1.4)<br>[n = 70] | 1.1 ± 1.6; 0.6 (0.4–0.8)<br>[n = 21] |
| Ferritin (ng/mL; normal range 23.9–336.2) | 760 ± 721; 622 (185–976)<br>[n = 66] | 693 ± 675; 567 (173–881)<br>[n = 56] | 1133 ± 883; 867 (522–1624)<br>[n = 10] |
| Severity-Related Biomarkers |  |  |  |
| Respiratory rate (breaths per minute) | 19 ± 3; 18 (18–19.5) | 18 ± 2; 18 (17–19) | 21 ± 5; 19 (18–20) |
| Ratio of PaO<sub>2</sub> to FiO<sub>2</sub>* | 259 ± 140; 200 (149–384)<br>[n = 88] | 277 ± 144; 230 (160–393)<br>[n = 67] | 201 ± 110; 142 (130–233)<br>[n = 21] |
| Pulse (beats per minute) | 86 ± 15; 85 (76–95) | 84 ± 15; 84 (72–90) | 96 ± 13; 95 (90–101) |
| Mean pressure (mmHg) | 86 ± 14; 86 (74–94)<br>[n = 92] | 87 ± 15; 87 (72–111)<br>[n = 70] | 83 ± 10; 84 (74–89) |
| Temperature (°C) | 36.9 ± 0.8; 36.5 (36.2–37.5) | 36.9 ± 0.8; 36.7 (36.2–37.5) | 36.8 ± 0.9; 36.4 (36.2–38) |
| SOFA* | 4 ± 3; 4 (1–6) | 3.5 ± 2.8; 3 (1–6) | 5.6 ± 2.7; 6 (4–7) |
| Comorbidities |  |  |  |
| Hypertension | 50 (52) | 39 (53) | 11 (50) |
| Chronic heart diseases | 14 (15) | 11 (15) | 3 (14) |
| Atrial fibrillation | 12 (13) | 10 (14) | 2 (9) |
| Diabetes type 2 | 29 (30) | 22 (30) | 7 (32) |
| Obesity (BMI) | 28.3 ± 7.7; 26 (23–31)<br>[n = 80] | 28.3 ± 8.3; 26 (23–33)<br>[n = 66] | 27.8 ± 4.0; 27.7 (24–29)<br>[n = 14] |
| Chronic obstructive pulmonary disease | 9 (9) | 7 (9) | 2 (9) |
| Chronic Kidney disease | 8 (8) | 6 (8) | 2 (9) |
| History of neoplasia | 14 (15) | 14 (19) | 0 (0) |
| Stroke | 9 (9) | 5 (7) | 4 (18) |

Wilcoxon rank-sum test and two-tailed Fisher's exact test between death and alive patients: \* $p < 0.05$ ; \*\* $p < 0.001$

In our cohort, 52 patients (54.2%) were transferred to the ICU within 7 days from admission because of clinical worsening. The primary endpoint of death occurred in 22 patients (23%), comprised of 14 patients who died in ICU and 8 in medical wards, while a total of 58 patients (60,4%) had been discharged and 16 (16,6%) were still hospitalized. PTX3 values were higher in dead patients compared to survivors (mean 52.3±41.8 ng/mL; median 39.8 ng/mL, IQT 20.2-75.7 ng/mL versus mean 21.0±20.7 ng/mL; median 15.3 ng/mL, IQT 8.2-21.3 ng/mL, respectively; Table 1 and Figure 3 panel A); moreover, higher PTX3 levels were also found in ICU patients compared to ward patients (mean 32.4±27.7 ng/mL; median 21.0 ng/mL, IQT 15.6-46.3 ng/mL versus mean 24.3±31.8 ng/mL; median 12.4 ng/mL, IQT 6.12-20.2 ng/mL, respectively; Figure 3 panel B).

**Figure 3.**
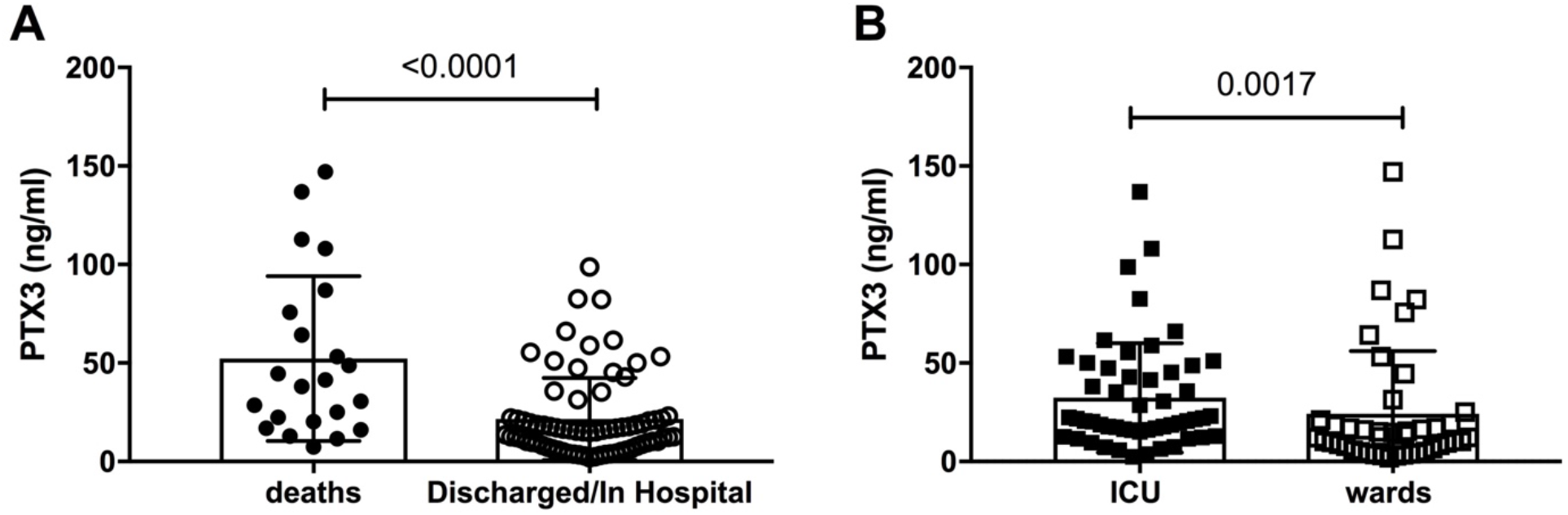
PTX3 plasma levels in 96 COVID-19 patients admitted to Humanitas Clinical and Research Hospital. Left panel: PTX3 plasma levels in patients based on the primary outcome (mortality). Right panel: PTX3 plasma levels in patients admitted to medical wards (50) or ICU (46).

The best AUC value for the prediction of death at 28 days was calculated for PTX3 (0.75, 95% CI 0.64–0.86). The most valid cut-off level to predict 28-day mortality was PTX3 = 22.25 ng/mL (sensitivity 0.73; specificity 0.78; Figure 4). The Kaplan-Meier curve showed an overall 28-day event-free survival of 0.94±0.03 (95% CI, 0.83 to 0.97) in low PTX3 group (<22.25 ng/mL) and 0.52±0.09 (95% CI, 0.34 to 0.67) in high PTX3 group (≥22.25 ng/mL; Log-ranks test p<0.0001; Figure 5). In the univariate COX analysis, PTX3 was a strong predictor of mortality in COVID-19 patients (unadjusted HR for ≥22.25 versus <22.25 ng/mL, 10.25; 95% CI, 3.41 to 30.75; p<0.0001). Basing on available literature and statistical relevance, we adjusted the model for possible confounding factors (age, ICU stay, and SOFA score at admission). PTX3 was confirmed as a strong predictor of short-term mortality (adjusted HR for ≥22.25 vs <22.25 ng/mL, 7.8; 95% CI, 2.5 to 24.0; p<0.0001; Table 2). PH assumption was not violated (global test of PH assumption, p= 0.153).

**Table 2.** PTX3 as predictor of death in Hospitalized COVID-19 patients (multivariate Cox model, HR adjusted for confounders: age, stay in ICU, and SOFA score at admission).

| Variables | HR | Std. Error | 95% CI | *p*-Value |
| --- | --- | --- | --- | --- |
| PTX3 ($\geq 22.25$ ng/mL) | 7.8 | 4.47 | (2.54 – 24.03) | <0.0001 |
| Age | 1.09 | 0.036 | (1.03 – 1.17) | 0.005 |
| ICU | 1.76 | 1.43 | (0.36– 8.61) | 0.485 |
| SOFA | 1.22 | 0.12 | (1.01 – 1.47) | 0.036 |

**Figure 4.**
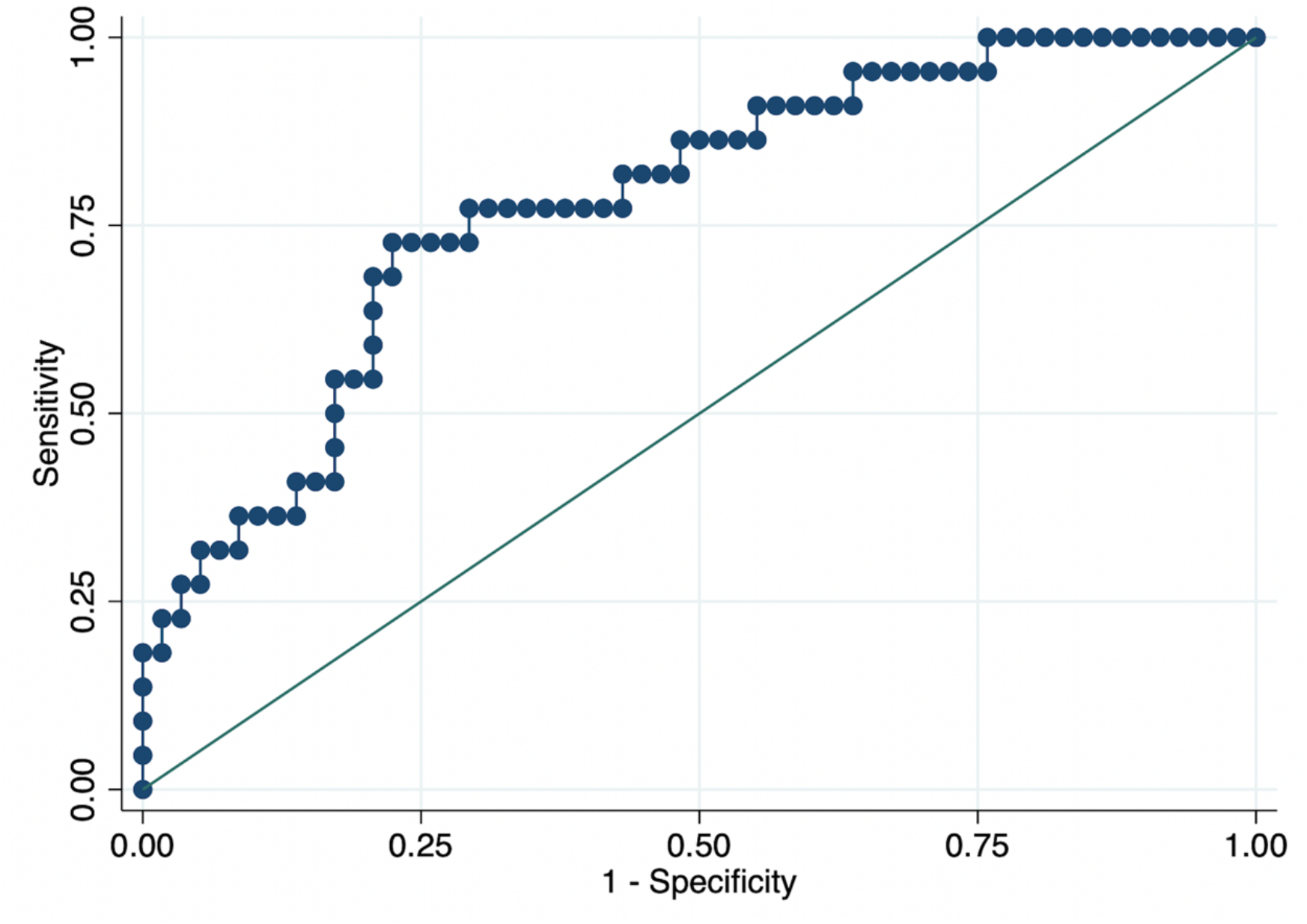
Receiver operating characteristic (ROC) curve for PTX3 for the primary outcome (death).

**Figure 5.**
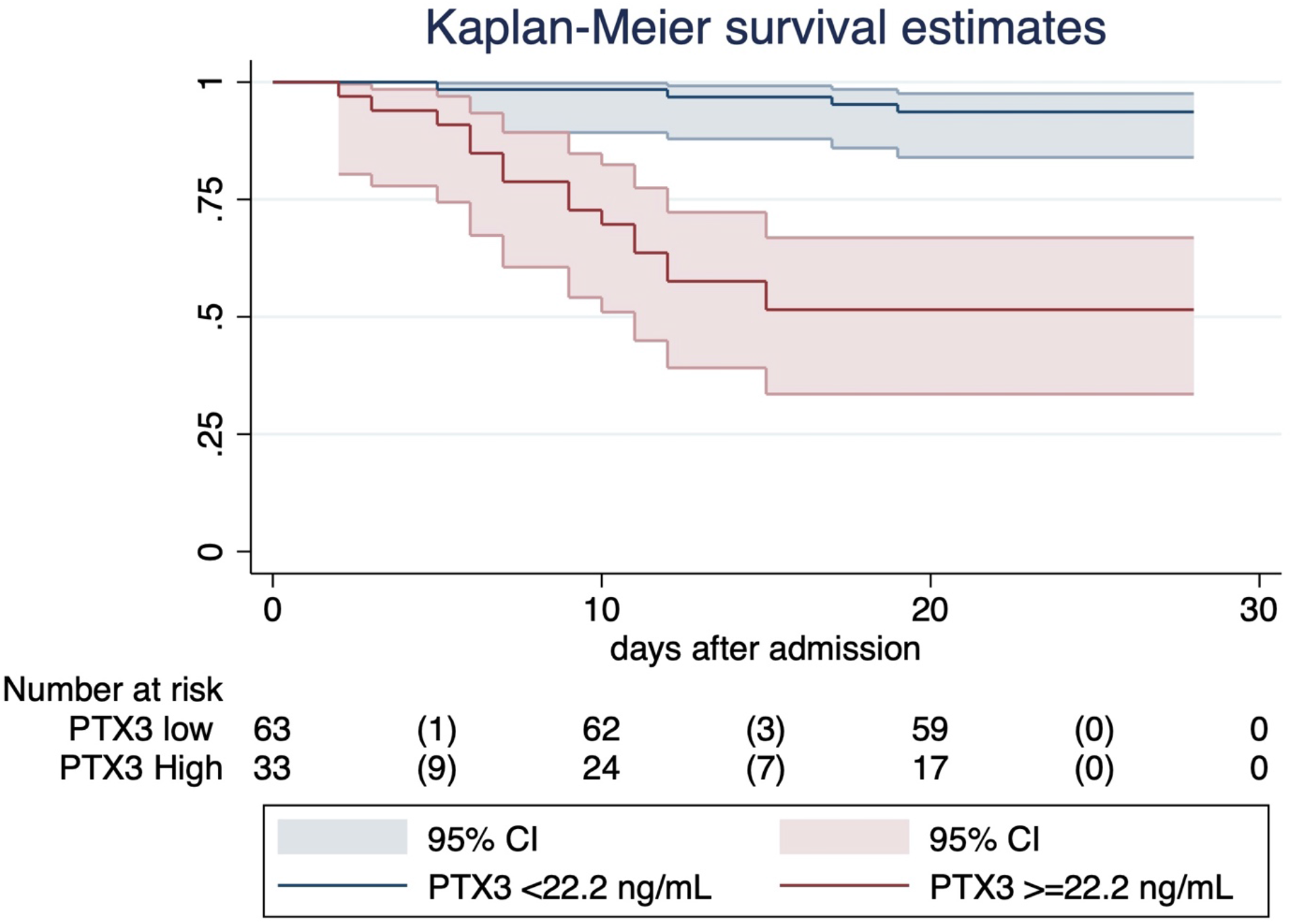
Kaplan-Meier curves by level of PTX3. High level was defined as ≥ of 22.2ng/mL, low level as < 22.2ng/mL. The 28-day event-free survival was 0.94±0.03 (95% CI, 0.83 to 0.97) in low PTX3 group and 0.52±0.08 (95% CI, 0.34 to 0.67) in high PTX3 group. CI: confidence interval.

Correlations between PTX3 and other inflammatory markers were initially assessed with Spearman test and were reported in Table 3. PTX3 was significantly correlated to CRP, procalcitonin, IL-6, ferritin, and D-dimer, but also with other COVID-19 poor prognostic factors, such as LDH, troponin-I, lymphocyte count (Table 3). PTX3, CRP, ferritin, IL-6, and D-dimer were also analyzed as continuous values for predicting mortality (univariate Cox regression; Table 4). PTX3 was confirmed to be significantly associated to mortality; moreover, IL-6 and D-dimer were also significantly, but weakly, associated with mortality at the unadjusted analysis. Performing a multivariate logistic regression including all these markers (PTX3, ferritin, D-Dimer, IL-6, and CRP), PTX3 was the only inflammatory marker significantly associated with death (adjusted HR for 1 ng/mL increase, 1.13; 95% CI, 1.02 to 1.24; p=0.021; Table 5).

**Table 3.** Spearman correlation between PTX3 and other inflammatory or biochemical markers.

| Variables | $\rho$ | *p*-Value |
| --- | --- | --- |
| Neutrophils | 0.0929 | 0.368 |
| Lymphocytes | -0.3048 | 0.002 |
| C-reactive protein | 0.3156 | 0.002 |
| Procalcitonin | 0.2841 | 0.006 |
| Interleukin-6 | 0.5353 | <0.001 |
| Ferritin | 0.4604 | <0.001 |
| D-dimer | 0.2945 | 0.017 |
| Lactate dehydrogenase | 0.5443 | <0.001 |
| Platelets | 0.0297 | 0.808 |
| Troponin-I | 0.5552 | <0.001 |

**Table 4.**
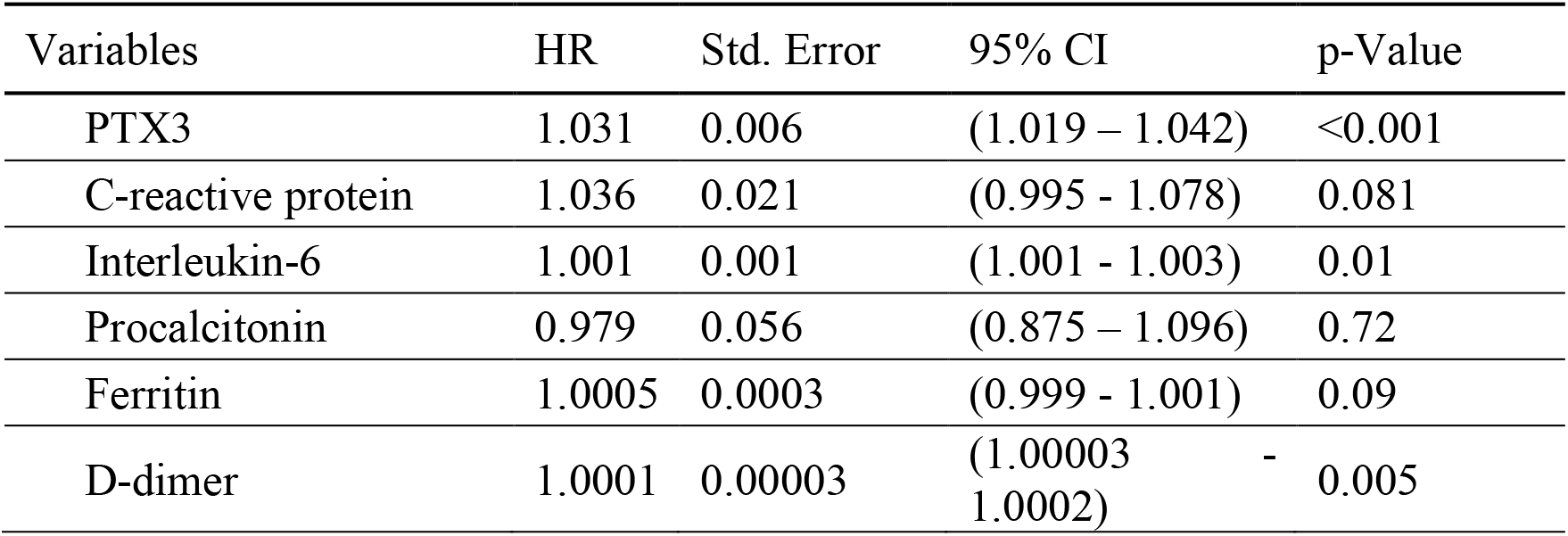
Inflammatory and other biomarkers as predictors of death in Hospitalized COVID-19 patients (univariate Cox models).

**Table 5.**
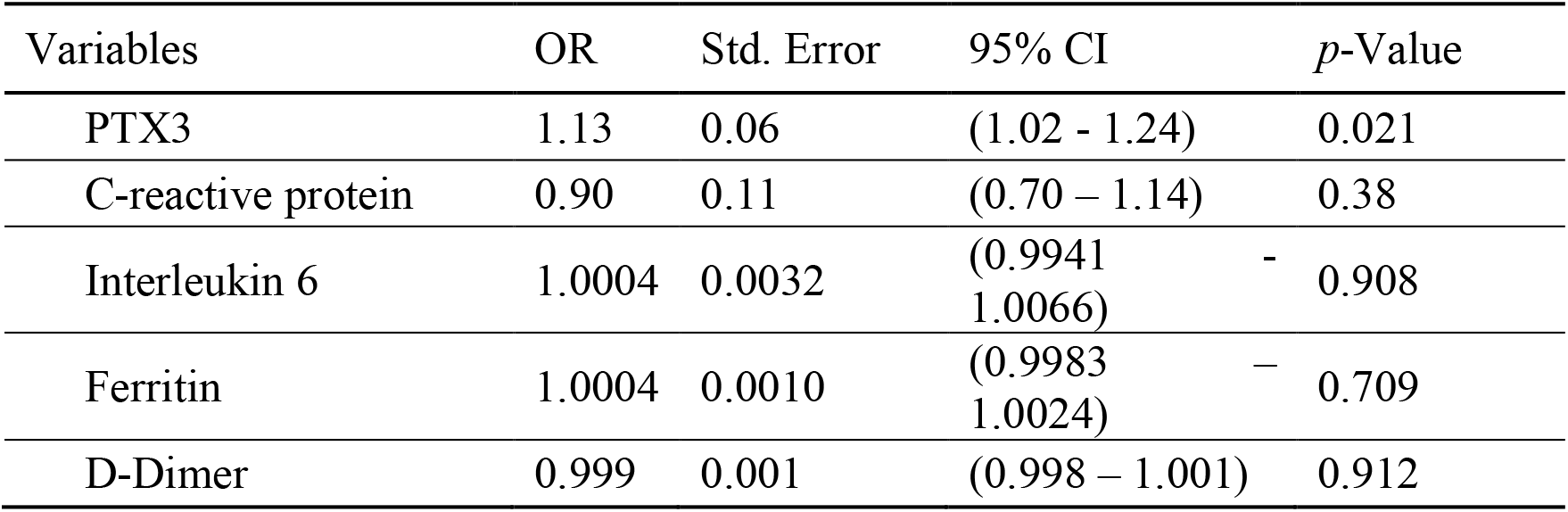
Correlations between inflammatory and other biomarkers levels (at admission) and outcome (death) in Hospitalized COVID-19 patients (multivariate logistic regression).

To investigate if PTX3 could be also considered a biomarker of COVID-19 severity at the time of hospital admission, identified by basal SOFA score ≥3, we performed a multivariate logistic regression including PTX3, CRP, IL-6, and D-dimer. CRP was the only factor significantly associated to SOFA score ≥3 (Table 6).

**Table 6.** Correlations between inflammatory and other biomarkers levels and systemic organ failure (SOFA score ≥ 3) at admission in Hospitalized COVID-19 patients (multivariate logistic regression).

| Variables | OR | Std. Error | 95% CI | *p*-Value |
| --- | --- | --- | --- | --- |
| PTX3 | 1.01 | 0.0128 | (0.98 – 1.03) | 0.508 |
| C-reactive protein | 1.14 | 0.0481 | (1.05 – 1.23) | 0.003 |
| Interleukin 6 | 1.00 | 0.0009 | (0.998 – 1.002) | 0.927 |
| D-Dimer | 1.00 | 0.0003 | (1.0 – 1.001) | 0.218 |

## DISCUSSION

As of May, 25^th^ the case fatality rate of COVID-19 in Italy is reported to be 14.3% with an average ICU admission rate of more than 20.4% ^45^. Elevated levels of CRP, cytokines, and chemokines ^8,44,46^ together with low lymphocyte and eosinophil counts characterize patients with severe disease ^47^. However, a reliable biomarker of poor outcome in COVID-19 is still lacking. The early and accurate triaging of the patients may contribute to better patient management and stratification in clinical trials. The present study was designed to investigate expression and clinical significance of the fluid phase pattern recognition receptor PTX3 in COVID-19. We found that PTX3 was induced by SARS-CoV-2 in respiratory tract epithelial cells. In COVID-19 patients, PTX3 analyzed at single cell level was selectively expressed by monocytes among circulating cells and by lung macrophages. High PTX3 plasma levels (≥ 22.25 ng/mL) were a strong independent indicator of short term 28-day mortality with an adjusted Hazard Ratio of 7.8 (95% CI 2.5-24). In this patient cohort, PTX3 fared definitely better than other known prognostic markers including CRP, IL-6, ferritin and D-dimer.

PTX3 blood levels above the normal value (<2 ng/mL ^48^) can be found in the subclinical inflammatory status of cardiovascular diseases ^49^ as well as in infections and sepsis, with increasing median values when moving to more severe conditions. In a large study conducted in 1326 unselected hospitalized subjects (14% with infectious diseases), PTX3 above 95th percentile of healthy non-hospitalized subjects (>6.4 ng/mL) was significantly associated to higher mortality in the short term, independently of hospitalization causes (adjusted HR 5, 95%CI 2.9-8.8) ^50^. Elevated PTX3 serum levels are indeed not related to a specific diagnosis rather predict severe cases or poor prognosis in different contexts characterized by a systemic inflammatory response ^50^. In a recent, prospective, observational study including 547 ICU patients (42.4% with infections), a PTX3 cut off similar to that identified in our study was reported to predict mortality: PTX3 serum level above the median cohort value of 20.9 ng/mL was independently associated to 28-day mortality when adjusted for age, sex, chronic diseases, and immunosuppression (HR 1.87, 95% CI 1.41-2.48) ^51^. In another recent paper conducted on 281 sepsis patients, serum PTX3 >26 ng/mL was associated to mortality ^52^. Taken together, these findings and our results suggest that circulating PTX3 levels ten-fold above the normal value reflect a severe systemic inflammatory involvement with ominous outcome.

PTX3 has been shown to be produced by diverse cell types including myelomonocytic cells, lung epithelial cells and endothelial cells. In the present study, we found that SARS-CoV-2 induced gene expression of PTX3 in respiratory tract epithelial cells. Peripheral blood mononuclear cells represent the easily accessible cellular source in patients. By bioinformatic analysis at single cell level, we found that PTX3 was selectively expressed by monocytes among circulating leukocytes. Moreover, in lung bronchoalveolar lavage fluid, single cell analysis revealed selective expression of PTX3 in neutrophils and macrophages, which play a major role in the pathogenesis of the disease ^3,4,53^.

The PTX3 gene was originally cloned in endothelial cells ^12^ and vascular cells are a major source of this component of humoral innate immunity, though their role could not be directly ascertained in the present study. Endothelial cells and the lung vascular bed have emerged as major determinant of COVID-19-associated microvascular thrombosis and disease pathogenesis ^9^. PTX3 plasma levels have been shown to correlate with severity of disease in various forms of vascular pathology including small vessel vasculitis, coronary heart disease, and Kawasaki disease ^30,49,54^. The latter observation raises the issue of its significance in the Kawasaki-like disease observed in children after COVID-19 (e.g.^55,56^). Of interest, our data show a significant correlation between PTX3 and D-dimer, surrogate of coagulation cascade activation and marker of venous thrombosis, and between PTX3 and troponin-I, marker of myocardial disease: both myocardial inflammation and acute ischemic heart disease have been described in COVID-19 ^57,58^. These observations raise the possibility that the strong prognostic significance of PTX3 in COVID-19 may reflect its positioning at the very intersection between macrophage-driven inflammation and vascular involvement.

PTX3 is a fluid phase pattern recognition molecule which binds selected viruses and play a role in anti-microbial immunity ^12^. Moreover, PTX3 has a regulatory role in inflammation by interfering with selectin-dependent neutrophils recruitment and by regulating the complement cascade ^59,60^. It is tempting to speculate that high levels of PTX3 in COVID-19 reflect failed negative regulation of uncontrolled inflammation. The actual role of PTX3 and, more in general, of humoral innate immunity in resistance against SARS-CoV-2 and in disease pathogenesis deserve further investigation.

In conclusion, the results presented here suggest that high PTX3 plasma levels (≥22 ng/mL) are strongly associated with unfavorable COVID-19 disease progression, defined as 28-day mortality, and may serve as a useful prognostic biomarker to decide intensity of care based on the predicted individual risk of death. PTX3 fared better than other classic biomarkers including CRP and IL-6. Given the relatively small sample size (96 patients) this finding should be interpreted with caution. With this caveat, it is tempting to speculate that PTX3 plasma levels may better reflect local tissue disruptive inflammation including the involvement of myelomonocytic cells and the vascular bed. The significance of PTX3 as a biomarker in COVID-19 patient management and stratification and its role in the virus-host interaction deserve further studies.

## Data Availability

The data that support the findings of this study are available from the corresponding author, [AM], upon reasonable request.

## Acknowledgements

We are grateful to Prof. Alessandro Rambaldi and Dr. Giovanni Gritti (Azienda Ospedaliera ASST Papa Giovanni XXIII, Bergamo, Italy) for helpful discussion. We also acknowledge the work and contribution of Monica Rimoldi, Paolo Tentorio and Sonia Valentino (Humanitas Clinical and Research Center, Rozzano, Milano, Italy) for their tireless work on the collection of biological samples from COVID-19 patients.

